# Plasma eMTBR-tau as a marker of clinical stage and progression across the Alzheimer’s disease continuum

**DOI:** 10.64898/2026.10.21.26366056

**Authors:** Johannes C. Michaelian, Rachael Yu, Alessandra Lee, Thomas K. Karikari, Lisa Shi, Eleanor Drummond, Sharon L. Naismith

## Abstract

**Importance:** Plasma biomarkers of neurodegeneration may help characterise progression across the Alzheimer’s disease (AD) continuum. Plasma eMTBR-tau243 has been associated with tau pathology and cognitive decline. Whether plasma eMTBR-tau levels differ across subjective cognitive decline (SCD), mild cognitive impairment (MCI) and AD dementia, between MCI subtypes, or best discriminate subsequent clinical progression in brain health and memory clinic populations remains unclear.

**Objective:** To compare plasma eMTBR-tau and selected biomarkers of neurodegeneration, neuropathology, and neuroinflammation across the AD continuum by pTau217 and *APOE* ε4 status, and explore their associations with clinical progression over two years.

**Design:** Observational cohort study with longitudinal follow-up in a subset of participants.

**Setting:** Healthy Brain Ageing (HBA) brain health and memory clinic, Sydney, Australia. Participants underwent assessment between October 2018 and April 2026.

**Participants:** Cross-sectional analysis included 436 participants; 166 were followed up approximately two years later.

**Exposures:** Plasma biomarker quantification using the NULISAseq^TM^ Neuro 220 Panel, clinical classification by multidisciplinary memory clinic assessment, plasma pTau217 status (SIMOA ALZpath), and *APOE* ε4 carrier status.

**Main Outcomes and Measures:** Fasting blood was collected on the morning of assessment. Plasma was analysed using the NULISAseq^TM^ Neuro 220 panel. Relative biomarker abundance was compared across clinical and biomarker-defined groups. BD-pTau217 and eMTBR-tau levels at the preceding assessment were modelled individually and jointly to assess SCD-to-MCI progression and MCI-to-AD conversion over approximately two years of follow-up.

**Results:** Among 436 participants (mean [SD] age, 70.2 [8.0] years; 67.2% female), 186 had SCD, 228 MCI, and 22 AD dementia. Among 221 biomarkers, phosphorylated tau analytes (pTau181/T205/T212/217/231) dominated clinical group differences, while eMTBR-tau showed the largest adjusted differences: 1.69-fold higher in MCI than SCD, 6.55-fold higher in AD than SCD and 3.88-fold higher in AD than MCI (all FDR-adjusted P < .001). eMTBR-tau was higher in multidomain amnestic MCI than in SCD, and in pTau217-positive than in pTau217-negative individuals. In the exploratory MCI-to-AD analysis (6/40 converted over 2 years), eMTBR-tau at the preceding assessment showed higher cross-validated discrimination of MCI-to-AD conversion than BD-pTau217 (AUC, 0.936 *vs.* 0.882).

**Conclusions and Relevance:** Clinical stages across the AD continuum are defined by a tau-dominant pattern. eMTBR-tau showed prominent stage-related differences, supporting its further evaluation as a marker of clinical progression.

**KEY POINTS:** *Question:* How do plasma biomarkers of neurodegeneration vary across clinical and biomarker-defined groups along the Alzheimer’s disease (AD) continuum, and are eMTBR-tau levels associated with clinical progression over approximately two years of follow-up?

*Findings:* In a brain health and memory clinic cohort of 436 participants, plasma eMTBR-tau showed the largest adjusted biomarker difference between subjective cognitive decline, mild cognitive impairment (MCI) and AD dementia groups. Amongst 166 participants followed up after 2 years, plasma eMTBR-tau levels at the preceding assessment best discriminated MCI-to-AD conversion (6/40; AUC=0.936).

*Meaning:* Plasma eMTBR-tau is a potential biomarker of clinical progression across the AD continuum.

## INTRODUCTION

Alzheimer’s disease (AD) is characterised by the accumulation of amyloid-β (Aβ) plaques and tau neurofibrillary tangles (NFTs) in the brain^1^. Importantly, the burden of tau NFT pathology is closely associated with clinical impairment and subsequent decline^2^. Hence, an important research and clinical goal^3^ is to establish robust, accessible methods for identifying and tracking tau NFT pathological burden across the AD continuum.

While plasma pTau217 is an established biomarker of AD-associated amyloid-β pathology^4^, it has been demonstrated to be elevated before tau NFT accumulation becomes detectable by positron emission tomography (PET)^5^. In this context, plasma endogenously cleaved microtubule-binding region tau (eMTBR-tau) has emerged as a marker of tau-PET burden and biological Braak stage, potentially complementing amyloid-β biomarkers by reflecting tau pathology across the AD continuum^3^. Together, both plasma pTau217 and eMTBR-tau may enable minimally invasive assessment of brain amyloid-β and tau NFT burden. While blood-based immunoassays for eMTBR-tau have recently become available^6^, it remains unclear how eMTBR-tau varies across clinical diagnostic groups and mild cognitive impairment (MCI) subtypes, and whether higher plasma eMTBR-tau levels at the preceding assessment predict subsequent clinical progression in brain health and memory clinic participants.

In this study, we investigated plasma eMTBR-tau as a marker of clinical stage and progression within a broader, data-driven analysis of plasma biomarkers measured using the NULISAseq™ Neuro 220 panel in participants attending the Healthy Brain Ageing (HBA) brain health and memory clinic for assessment. We first used cross-sectional volcano analyses to compare biomarkers reflecting neuropathology, neurodegeneration and neuroinflammation across subjective cognitive decline (SCD), mild cognitive impairment (MCI) and AD dementia groups, and between MCI subtypes. Secondarily, we examined differences between plasma pTau217-positive and -negative groups, and by *APOE* ε4 carrier status across the AD continuum. Lastly, we focused on eMTBR-tau and compared it with BD-pTau217, evaluating levels at the preceding assessment individually and jointly in exploratory, covariate-adjusted, cross-validated ROC analyses to assess discrimination of subsequent progression from SCD to MCI and conversion from MCI to AD over approximately two years of follow-up.

## METHODS

### Study design and participants

This observational study used data from participants assessed through the Healthy Brain Ageing (HBA) clinic, a specialist research brain health and memory clinic in Sydney, Australia. Requiring a GP or medical specialist referral, the clinic provides comprehensive neuropsychological and medical assessments for older adults aged ≥ 50 years. Exclusion criteria for participation at the HBA clinic include head injury with loss of consciousness > 30 min, neurological disease (e.g., Parkinson’s disease) or non-affective psychiatric illness (e.g., schizophrenia), intellectual disability, insufficient English for neuropsychological assessment, current or previous substance misuse, MMSE < 20 at the time of referral and/or a previously established diagnosis of dementia.

The present study included 436 unique participants, of whom 166 returned for a follow-up assessment after approximately two years. This yielded 602 participant-visit observations, each with a plasma sample available for NULISAseq^TM^ Neuro 220 Panel analysis.

### Memory clinic assessment and clinical classification

Subjective cognitive decline was defined as self-or informant-reported cognitive concerns prompting referral to the HBA clinic, without meeting criteria for MCI. MCI was classified by multidisciplinary consensus according to Winblad et al. (2004)^7^, with objective impairment defined as performance ≥1.5 SD below age-and education-adjusted normative expectations. MCI was subclassified as amnestic (aMCI) or non-amnestic (naMCI), and as single-domain (sd-) or multidomain (md-) according to the number of affected cognitive domains^8^. AD dementia was diagnosed according to clinical criteria for possible or probable AD, requiring cognitive impairment and functional decline that interfered with daily activities^9^.

#### Covariates relevant to plasma biomarker concentrations

As part of the medical assessment, participants were provided a referral for a fasting clinical blood test at a commercial Australian pathology collection centre (e.g., Douglass Hanly Moir, Australia). Based on the pathology report and relevant to this study, the estimated glomerular filtration rate (eGFR) was calculated using the CKD-EPI creatinine equation^10^. Contemporaneous eGFR was available for 543 of 602 plasma sample records (90.2%). To minimise exclusions from adjusted analyses, eGFR for 36 records (6.0%) was assigned from measurements at earlier or later visits or estimated from serial clinical results provided by the participant’s referring GP or medical specialist at the time of assessment. Values for the remaining 23 records (3.8%), for which no usable numerical eGFR was available, using predictive mean matching with the mice package in R^11^. Fifty completed datasets were generated for each variable, using 20 iterations and a pool of five candidate donor records for each imputed value. Body mass index (BMI) was calculated during assessment as weight in kilograms divided by height in metres squared, with contemporaneous values available for 556 of 602 records (92.4%). Missing BMI for 26 records (4.3%) was assigned from measurements at another visit for the same participant, while values for the remaining 20 records (3.3%), for which no usable BMI was available at any timepoint, were imputed using the same predictive mean matching approach described for eGFR. Completed eGFR and BMI values were treated as fixed covariates in a single analysis dataset.

### Blood collection

Fasting venous blood was collected on the morning (between 8:30 and 9:15 AM) of clinical assessment into ethylenediaminetetraacetic acid (EDTA) vacuum tubes. Within 30 minutes of collection, samples were centrifuged at 1,800 × g for 10 minutes at 4 °C. Plasma was aliquoted into cryovials and immediately stored at −80 °C until analysis.

### Biomarker analysis

At the time of Nucleic-acid Linked Immuno-Sandwich Assay (NULISA) and Single Molecule Array (SIMOA) analyses, all plasma samples underwent one freeze-thaw cycle in total (i.e., thawed for the first time since freezing).

#### NULISAseq^TM^ Neuro 220 Panel analysis

Plasma biomarker profiling was performed through Panomics Biosciences Pty Ltd, an Alamar Biosciences Certified Service Provider, by GeneWorks Pty Ltd in Australia using the NULISAseq™ Neuro 220 Panel (Alamar Biosciences, Fremont, California, USA). Sample analysis and data processing were performed according to the manufacturer’s instructions, and as described previously^12^. Relevant to this study, all 602 plasma samples were randomised across seven assay plates and analysed undiluted in singlicate using the same assay lot (2614702)^13^ and instrument (serial number 10073). Data were exported using NULISA Analysis Software (NAS), version 1.5.3, and reported as NULISA Protein Quantification (NPQ) values, representing normalised relative biomarker abundance on a log2 scale.

#### SIMOA plasma pTau217 analysis

Plasma concentrations of pTau217 (n=354; 59% of the 602 samples with NULISA data) were quantified on the Quanterix SIMOA HD-X platform (Quanterix, Billerica, MA, USA). Plasma pTau217 concentration was measured using the SIMOA ALZpath pTau217 kit (lot #504400). Across all plates, samples were run according to the manufacturer’s instructions using the 4x dilution protocol, with values automatically adjusted for dilution by the instrument. All samples were assayed in duplicate; if the CV exceeded 20%, measurements were repeated in duplicate, and the final value was derived from 3 of 4 replicates. Participants were classified as pTau217-negative (plasma pTau217 concentration <0.4 pg/mL) or pTau217-positive (plasma pTau217 concentration ≥0.64 pg/mL) based on reference ranges providing 95% sensitivity and specificity in detecting amyloid-PET positivity^4^. Participants with pTau217 concentrations in between the cut-off ranges (0.4-0.64 pg/mL) were considered indeterminate and not included in analyses stratified by pTau217 status.

To estimate SIMOA pTau217 concentrations where direct measurements were unavailable, we fitted a linear model relating log₂ SIMOA pTau217 to NULISA pTau217 NPQ in 354 paired observations from 277 participants. Predictions were converted to pg/mL using a Duan smearing correction, yielding estimated SIMOA pTau217=0.00060747 × 2^(1.00560014 × NULISA pTau217 NPQ)^. In 10-fold cross-validation grouped by participants, predicted and observed concentrations were strongly correlated (*r*=0.932; predictive R²=0.866), with a mean absolute error of 0.085 pg/mL and 94.4% coverage of the 95% prediction intervals. Of 248 samples without direct SIMOA measurements, two fell outside the calibration range and were excluded, leaving 246 estimated concentrations for analysis. Observed SIMOA values were retained where originally available.

### Apolipoprotein E (*APOE)* ε4 status

Although formal *APOE* genotyping was available for 306 of 436 participants (70.2%), across cross-sectional and longitudinal observations, *APOE* ε4 carrier status in the analyses was inferred from APOE4 protein expression measured by the NULISA Neuro 220 Panel. APOE4 protein NPQ values formed two clearly separated clusters, with a maximum of 6.68 in the low-abundance cluster and a minimum of 11.34 in the high-abundance cluster. An NPQ cut-off of 11.34, corresponding to the lower boundary of the high-abundance cluster, was used to classify samples as *APOE* ε4 carriers (NPQ ≥ 11.34) or non-carriers (NPQ < 11.34). Among samples with formal genotyping (n=466), the APOE4 protein-based NPQ classification agreed with genotype-derived *APOE* ε4 status in 98.7% of cases.

### Statistical analysis

HBA participant characteristics were summarised as mean (standard deviation) for continuous variables and frequency (percentage) for categorical variables. Between-group differences were assessed using one-way analysis of variance, Kruskal-Wallis or χ² tests, as appropriate. Significant analysis of variance and Kruskal-Wallis results were followed by pairwise t tests and Dunn tests, respectively^14^. For cross-sectional proteomic analyses, each participant contributed one sample, prioritising their latest available assessment. Differences in log₂-scaled NPQ values were assessed using empirical Bayes moderated linear models, with adjustment for age, sex, *APOE* ε4 carrier status, eGFR, BMI and assay plate as appropriate to each comparison^15,16^. Results were expressed as adjusted log₂ fold changes. Biomarker-level *p* values were corrected using the Benjamini-Hochberg method within each comparison, with a false discovery rate < 0.05 considered significant^17^. The ability of BD-pTau217 and eMTBR-tau levels at the preceding assessment to distinguish subsequent SCD-to-MCI progression and MCI-to-AD conversion was assessed using receiver operating characteristic (ROC) curves and areas under the curve (AUCs) derived from stratified, threefold cross-validated ridge logistic regression models^18,19^. Models evaluated each biomarker individually and both jointly, with adjustment for age, sex, eGFR, BMI and assay plate. Ridge regularisation, with a fixed penalty of λ=0.1, was used to limit coefficient instability and overfitting. Threefold stratification accommodated the small number of progression events while retaining converters and non-converters in each validation fold. Cross-validation was performed once using a fixed random seed, and AUCs were calculated from pooled held-out predictions. 95% confidence intervals were estimated using 1,000 stratified bootstrap resamples conditional on these predictions; stability across alternative fold assignments was not assessed. For both endpoints, progression status was determined from the clinical diagnosis at the most recent follow-up assessment. Follow-up duration was calculated as the interval between the preceding and most recent assessments, averaging approximately two years. All tests were two-sided, with *p* < 0.05 considered significant unless otherwise specified. Analyses and visualisations were performed in R using RStudio.

### Data approval statement

All participants provided written consent prior to study participation, and all research activities were approved by the University of Sydney Human Research Ethics Committee (Protocol numbers: 2012/1873 and 2019/HE000271). All study methods were conducted in compliance with the Helsinki Declaration.

## RESULTS

### Cohort characteristics

**Table 1** reports on participant characteristics at their latest clinical assessment, which was also used for the primary cross-sectional analyses. The cohort comprised 436 participants including 186 with SCD, 228 with MCI, and 22 with AD dementia. The mean age of participants was 70.2 years, with those in the MCI (mean=71.6 years) and AD (mean=74.2 years) groups significantly older than those with SCD (mean=68.1 years). Approximately two-thirds were female (n=293, 67.2%), and 39.7% (n=173) were *APOE* ε4 carriers. As expected, MMSE scores were highest in the SCD group (mean=29.4), followed by MCI (mean=27.8) and AD (mean=23.4), with all pairwise differences significant (*p* < 0.001). Mean plasma pTau217 concentrations were lowest in SCD (0.32 pg/mL), followed by MCI (0.61 pg/mL) and AD (0.92 pg/mL), with significant differences between each group. eGFR also differed across groups in the overall test (*p*=0.034), with significantly lower levels in AD than in SCD (*p*=0.026) and MCI (*p=*0.041).

**Table 1.** Characteristics of the HBA study cohort at the latest sampled assessment, overall and by clinical diagnostic group.

| Characteristic | Overall<br>(n=436) | SCD<br>(n=186) | MCI<br>(n=228) | AD<br>(n=22) | Omnibus<br>*p* | Significant pairwise<br>comparisons |
| --- | --- | --- | --- | --- | --- | --- |
| Age (years) | 70.2 (8.0) | 68.1 (7.3) | 71.6 (8.1) | 74.2 (7.5) | <0.001 | MCI > SCD ($p<0.001$);<br>AD > SCD ($p=0.003$) |
| Sex, female | 293 (67.2%) | 130 (69.9%) | 149 (65.4%) | 14 (63.6%) | 0.579 | None |
| Education (years) | 14.7 (3.0) | 14.8 (2.9) | 14.6 (3.1) | 14.6 (3.5) | 0.689 | None |
| MMSE | 28.3 (2.3) | 29.4 (0.8) | 27.8 (2.4) | 23.4 (2.5) | <0.001 | SCD > MCI ($p<0.001$);<br>SCD > AD ($p<0.001$);<br>MCI > AD ($p<0.001$) |
| eGFR (mL/min/1.73 m<sup>2</sup>) | 79.8 (10.9) | 80.9 (9.7) | 79.6 (11.3) | 72.8 (13.5) | 0.034 | SCD > AD ($p=0.026$);<br>MCI > AD ($p=0.041$) |
| BMI (kg/m<sup>2</sup>) | 26.0 (4.6) | 26.1 (4.5) | 25.9 (4.7) | 27.2 (5.0) | 0.486 | None |
| *APOE* ε4 carriers | 173 (39.7%) | 66 (35.5%) | 95 (41.7%) | 12 (54.5%) | 0.151 | None |
| Plasma pTau217 (pg/mL) | 0.50 (0.407) | 0.32 (0.198) | 0.61 (0.457) | 0.92 (0.480) | <0.001 | MCI > SCD ($p<0.001$);<br>AD > SCD ($p<0.001$);<br>AD > MCI ($p=0.004$) |
| Plasma pTau217 status<sup>1</sup> | | | | | <0.001 | MCI vs. SCD distributions<br>differ ($p<0.001$);<br>AD vs. SCD distributions<br>differ ($p<0.001$);<br>AD vs. MCI distributions<br>differ ($p=0.009$) |
| Negative (<0.4 pg/mL) | 256 (59.0%) | 145 (78.0%) | 108 (47.4%) | 3 (15.0%) |  |  |
| Indeterminate (0.40 to <0.64<br>pg/mL) | 60 (13.8%) | 29 (15.6%) | 28 (12.3%) | 3 (15.0%) |  |  |
| Positive ($\geq 0.64$ pg/mL) | 118 (27.2%) | 12 (6.5%) | 92 (40.4%) | 14 (70.0%) | | |

Each participant contributes their latest sampled assessment. Education uses the highest valid recorded value; BMI and eGFR use the existing completed values in the selected record.
Abbreviations: MMSE, Mini-Mental State Examination; eGFR, estimated glomerular filtration rate; BMI, body mass index; *APOE*, apolipoprotein E; MCI, mild cognitive impairment; SCD, subjective cognitive decline; AD, Alzheimer's disease.
<sup>1</sup> Study pTau217 thresholds: negative <0.4; indeterminate 0.40 to <0.64; positive $\geq 0.64$ pg/mL, using the combined measured/converted concentration. Thresholds are based on the SIMOA ALZpath references reported by Ashton et al (2024). Status comparisons assess the full three-category distribution.

### Plasma biomarker differences across the AD continuum

**Figure 1** illustrates differences in plasma biomarker abundance between SCD, MCI and AD groups, using each participant’s latest available sample. It shows that 17 biomarkers differed significantly between MCI and SCD, 22 between AD and SCD, and 17 between AD and MCI (FDR-adjusted *p* < 0.05) (see **eTable1 in the Supplement**). Of these, 16 were higher and one lower in MCI than SCD, while all significant biomarkers were higher in AD for both AD comparisons. Plasma eMTBR-tau was significantly higher in more advanced clinical groups and showed the largest adjusted fold change among significantly different biomarkers in each contrast. Specifically, plasma eMTBR-tau was 1.69-fold higher in MCI than SCD (95% CI, 1.38 to 2.06; FDR-adjusted *p*=1.03 × 10^-5^), 6.55-fold higher in AD than SCD (95% CI, 4.17 to 10.29; FDR-adjusted *p*=1.41 × 10^-13^), and 3.88-fold higher in AD than MCI (95% CI, 2.51 to 6.02; FDR-adjusted *p*=2.80 × 10^-7^).

**Figure 1.**
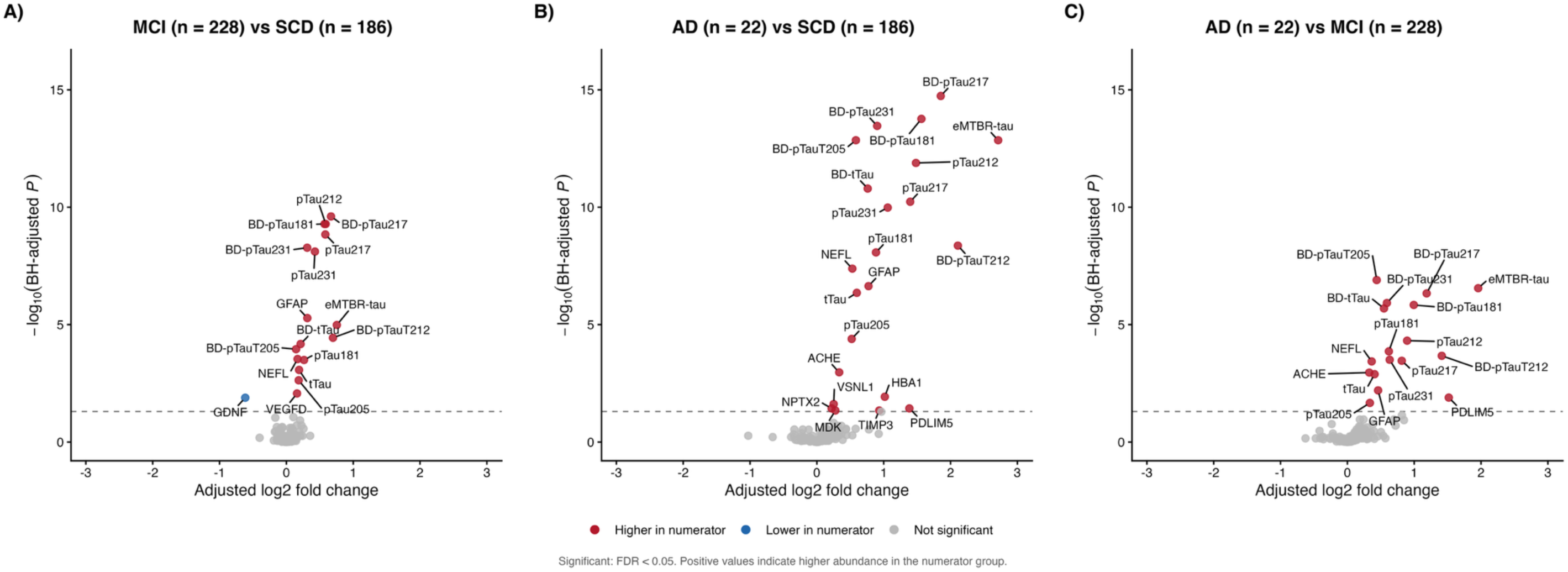
NULISAseq^TM^ Neuro 220 panel biomarker differences across the AD continuum Volcano plots compare **A)** MCI (n=228) with SCD (n=186), **B)** AD (n=22) with SCD, and **C)** AD with MCI (right). Analyses included one sample per participant from the latest eligible assessment. Empirical Bayes moderated linear models were adjusted for age, sex, *APOE* ε4 carrier status, eGFR, BMI and assay plate. Each point represents a biomarker. The x-axis shows the adjusted log₂ fold change, with positive values indicating higher abundance in the first named group; the y-axis shows −log₁₀ of the Benjamini-Hochberg-adjusted P value. Red and blue points indicate significantly higher and lower abundance, respectively; grey points indicate no significant difference. The dashed horizontal line marks the false discovery rate threshold of 0.05. Biomarkers meeting FDR < 0.05 are labelled. Abbreviations: AD, Alzheimer’s disease; *APOE*, apolipoprotein E; BMI, body mass index; eGFR, estimated glomerular filtration rate; FDR, false discovery rate; MCI, mild cognitive impairment; SCD, subjective cognitive decline.

Beyond eMTBR-tau, the significantly altered plasma biomarkers with the largest covariate-adjusted increases in abundance for MCI *vs.* SCD were BD-pTauT212, BD-pTau217 and pTau212; for AD *vs.* SCD were BD-pTauT212, BD-pTau217 and BD-pTau181; and for AD *vs.* MCI were PDLIM5, BD-pTauT212 and BD-pTau217. Full fold changes, 95% CIs and FDR-adjusted p values for all contrasts are reported in **eTable1 in the Supplement.**

### Plasma biomarker differences across SCD, md-aMCI and AD

Restricting the MCI group to md-aMCI (n=123) to reduce clinical heterogeneity, tau-related differences in comparison to SCD and AD were more pronounced (**Supplementary Figure S1**). In particular, plasma eMTBR-tau was 2.48-fold higher in md-aMCI than in SCD (95% CI, 1.94 to 3.16; FDR-adjusted p=6.88 × 10^-11^), compared with 1.69-fold higher levels for the overall MCI group relative to SCD. Plasma eMTBR-tau remained significantly higher in AD than in md-aMCI, with an adjusted fold change of 2.87 (95% CI, 1.81 to 4.54; FDR-adjusted *p*=9.90 × 10^-4^).

Beyond eMTBR-tau, the largest positive adjusted fold changes for md-aMCI *vs.* SCD were: BD-pTau217, BD-pTauT212 and pTau217 (similar to MCI *vs.* SCD contrasts); and PDLIM5, BD-pTauT212, HBA1 for AD *vs.* md-aMCI. Fold changes, 95% CIs and FDR-adjusted p values for all contrasts are reported in **eTable2 in the Supplement.**

### Plasma biomarker differences across MCI subtypes

**Figure 2** reports on all six pairwise comparisons among sd-aMCI (n=18), md-aMCI (n=123), sd-naMCI (n=53), and md-naMCI (n=34). Following covariate adjustment and FDR correction, significant differences were only observed between sd-naMCI and md-aMCI, with 13 biomarkers lower in the sd-naMCI group (**see eTable3 in the Supplement**). Plasma eMTBR-tau showed the largest proportional decrease among the significantly decreased biomarkers (adjusted fold change=0.40; 95% CI, 0.28 to 0.57; FDR-adjusted p=4.18 × 10^-5^), corresponding to approximately 60% lower abundance in sd-naMCI than in md-aMCI. Full fold changes for all other significant biomarkers contrasting within and between all MCI subtypes, 95% CIs and FDR-adjusted p values for all contrasts are reported in **eTable3 in the Supplement.**

**Figure 2.**
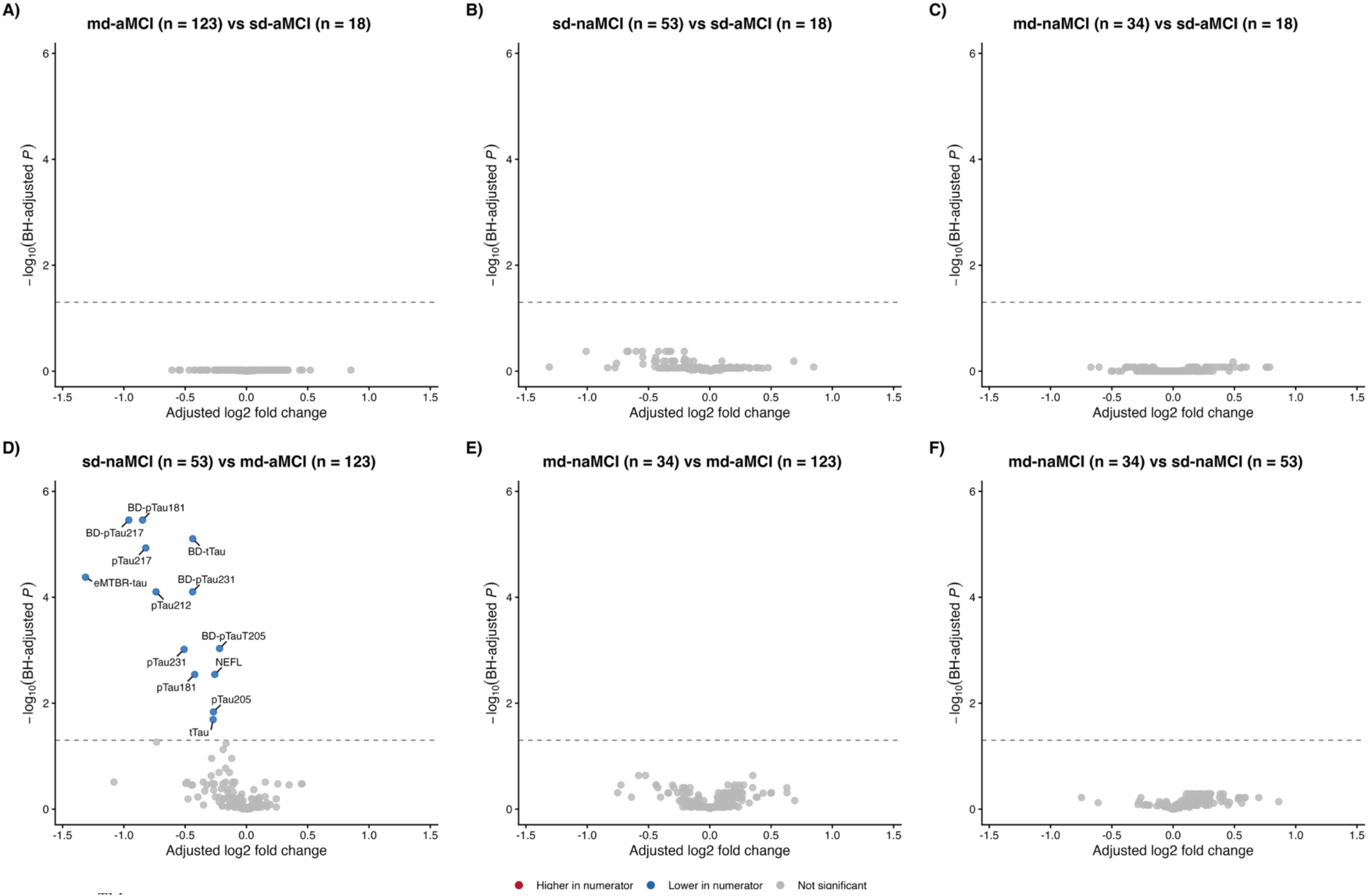
NULISAseq^TM^ Neuro 220 panel biomarker differences between MCI subtypes Volcano plots show all six pairwise comparisons among single-domain amnestic MCI (sd-aMCI; n=18), multidomain amnestic MCI (md-aMCI; n=123), single-domain non-amnestic MCI (sd-naMCI; n=53), and multidomain non-amnestic MCI (md-naMCI; n=34). Panels compare **A)** md-aMCI *vs.* sd-aMCI; **B)** sd-naMCI *vs.* sd-aMCI; **C)** md-naMCI *vs.* sd-aMCI; **D)** sd-naMCI *vs.* md-aMCI; **E)** md-naMCI *vs.* md-aMCI; and **F)** md-naMCI *vs.* sd-naMCI. Analyses used one sample per participant from the latest eligible assessment and empirical Bayes moderated linear models adjusted for age, sex, *APOE* ε4 carrier status, eGFR, BMI and assay plate. P values were adjusted across tested biomarkers separately within each comparison. Each point represents a biomarker. The x-axis shows the adjusted log₂ fold change and the y-axis shows - log₁₀ of the Benjamini-Hochberg-adjusted P value. Positive values indicate higher abundance in the first named group within each panel. Red and blue points indicate significantly higher and lower abundance, respectively; grey points indicate no significant difference. The dashed horizontal line marks FDR=0.05, with significance defined as FDR < 0.05. Significantly different biomarkers are labelled. Abbreviations: *APOE*, apolipoprotein E; BMI, body mass index; eGFR, estimated glomerular filtration rate; FDR, false discovery rate; MCI, mild cognitive impairment.

### Plasma biomarker differences by pTau217 status

Comparison of pTau217-positive (n=118) and pTau217-negative participants (n=256) identified 24 biomarkers with significantly different abundance after covariate adjustment and FDR correction (**Figure 3**). Of these, 22 were higher (predominantly tau biomarkers) and two lower in pTau217-positive participants (Aβ42 and APOA1). BD-pTau217 showed the largest adjusted fold change (4.47-fold higher), as expected given the pTau217-based grouping, closely followed by eMTBR-tau, which was 4.42-fold higher in pTau217-positive than in pTau217-negative participants (95% CI, 3.54 to 5.51; FDR-adjusted p=2.19 × 10^-31^). Full fold changes for all other significant biomarkers, 95% CIs and FDR-adjusted p values for all contrasts are reported in **eTable4 in the Supplement**.

**Figure 3.**
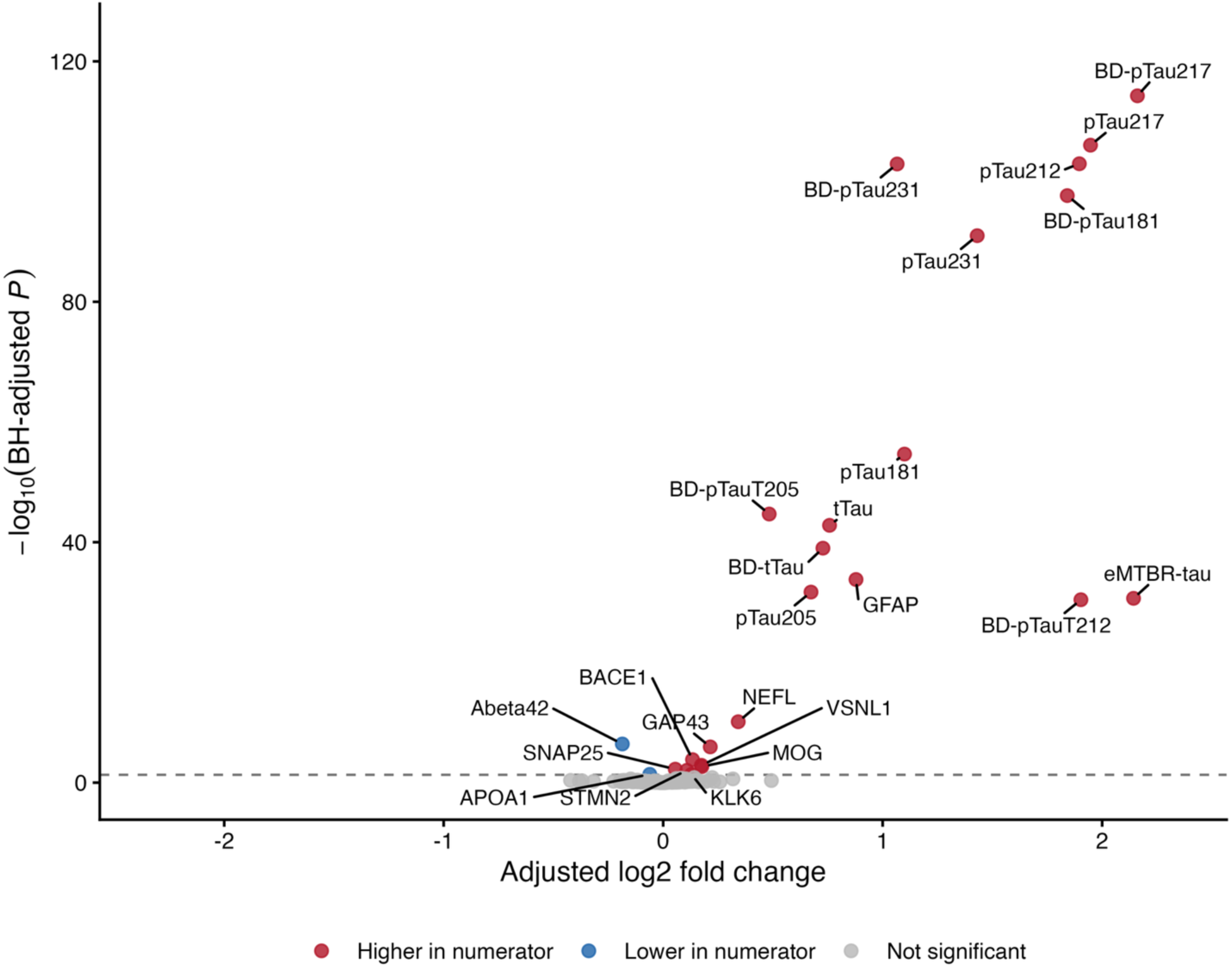
NULISAseq^TM^ Neuro 220 panel biomarker differences between pTau217-positive (n=118)^1^ and pTau217-negative (n=256)^1^ participants The volcano plot compares pTau217-positive (n=118) and pTau217-negative (n=256) participants. Positive status was defined as plasma pTau217 ≥0.64 pg/mL and negative status as <0.40 pg/mL; participants with intermediate values (0.40 to <0.64 pg/mL) were excluded from this comparison. Analyses used one sample per participant from the latest eligible assessment and empirical Bayes moderated linear models adjusted for age, sex, *APOE* ε4 carrier status, eGFR, BMI and assay plate. P values were adjusted across tested biomarkers separately within each comparison. Each point represents a biomarker. The x-axis shows the adjusted log₂ fold change and the y-axis shows −log₁₀ of the Benjamini-Hochberg-adjusted P value. Positive values indicate higher abundance in the pTau217-positive group. Red and blue points indicate significantly higher and lower abundance, respectively; grey points indicate no significant difference. The dashed horizontal line marks FDR=0.05, with significance defined as FDR < 0.05. Significantly different biomarkers are labelled. ¹ Study pTau217 thresholds: negative <0.4; indeterminate 0.40 to <0.64; positive ≥0.64 pg/mL, using the combined measured/converted concentration. Thresholds are based on the SIMOA ALZpath references reported by Ashton et al (2024). Abbreviations: *APOE*, apolipoprotein E; BMI, body mass index; eGFR, estimated glomerular filtration rate; FDR, false discovery rate; pTau217, tau phosphorylated at threonine 217.

### Plasma biomarker differences by *APOE* ε4 carrier status

11 biomarkers differed significantly within SCD and nine within MCI (**Supplementary Figure S2**). No biomarkers met the FDR-adjusted significance threshold within the AD group, which may reflect limited statistical power given the small sample size (n=22). Within SCD, nine significant biomarkers were higher (e.g., BD-pTau217, pTau217, BD-pTau181) and two lower in carriers (i.e., CRP and APOE); within MCI, seven were higher (e.g., BD-pTau217, pTau217, pTau212) and two lower (i.e., Aβ42 and APOE). However, eMTBR-tau did not differ significantly by carrier status within SCD (adjusted log2FC=0.25; 95% CI, −0.11 to 0.60; FDR-adjusted *p*=0.975), MCI (adjusted log2FC=0.33; 95% CI, −0.10 to 0.76; FDR-adjusted *p*=0.724), or AD (adjusted log2FC=-1.48; 95% CI, −3.24 to 0.28; FDR-adjusted *p*=0.610). Full fold changes for all other significant biomarkers, 95% CIs and FDR-adjusted p values are reported in **eTable5 in the Supplement**.

### Participant characteristics and exploratory ROC analyses of progression from SCD to MCI

A total of 106 participants with SCD at their preceding assessment timepoint were assessed for progression from SCD to MCI, of whom 19 (17.9%) converted to MCI during a mean follow-up of 2.0 years (**Supplementary Table S1**). At the preceding assessment, participants had a mean age of 67.1 years, 70.8% were female (n=75), and the mean MMSE score was 29.4. Participants who progressed to MCI and participants who remained SCD had similar mean ages (67.6 *vs.* 67.0 years), with a mean follow-up of 2.0 years in both groups. Mean MMSE scores at the preceding assessment were 29.7 *vs.* 29.3, and mean plasma pTau217 concentrations were 0.26 *vs.* 0.27 pg/mL, respectively.

Based on the cross-sectional findings, exploratory cross-validated ROC analyses focused on BD-pTau217 and eMTBR-tau, individually and in combination, to assess discrimination between participants who progressed to MCI and those who remained SCD over a mean follow-up of 2.0 years (**Figure 4A**). Following adjustment for age, sex, eGFR, BMI and assay plate, cross-validated AUCs were 0.477 (95% CI, 0.349 to 0.608) for BD-pTau217, 0.581 (95% CI, 0.442 to 0.718) for eMTBR-tau and 0.490 (95% CI, 0.356 to 0.635) for the model combining both BD-pTau217 and eMTBR-tau. At a fixed predicted-probability threshold of 0.50, all three models classified every participant as a non-converter (sensitivity, 0%; specificity, 100%; F1, 0).

**Figure 4.**
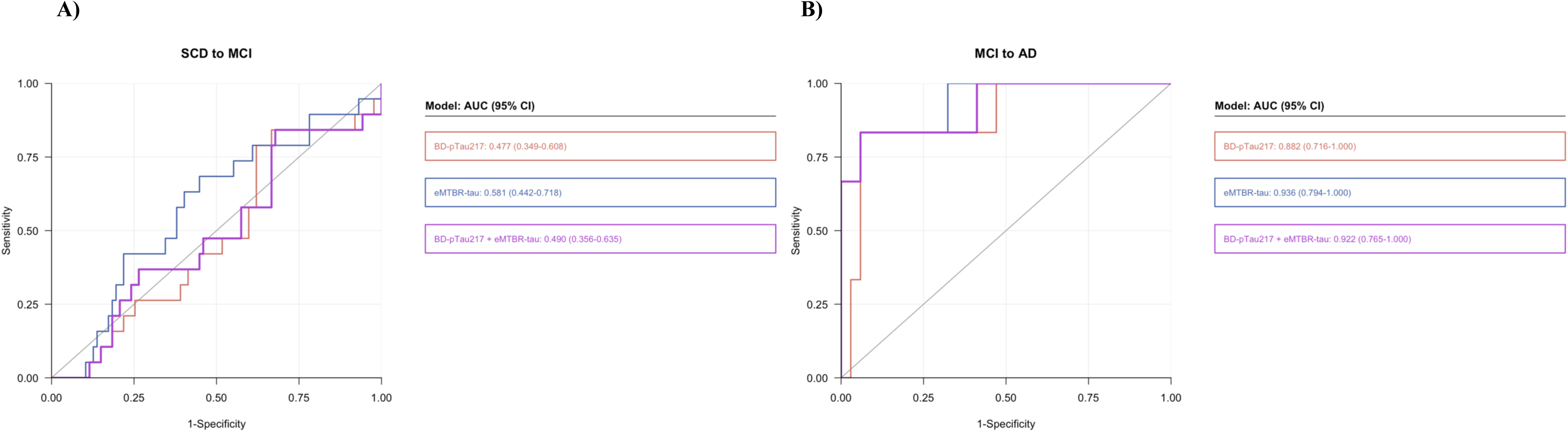
Exploratory cross-validated ROC analyses of (A) SCD-to-MCI progression and (B) MCI-to-AD conversion using plasma BD-pTau217 and eMTBR-tau. Receiver operating characteristic (ROC) curves show discrimination of (A) SCD-to-MCI progression (106 participants; 19 progressors) and (B) MCI-to-AD conversion (40 participants; 6 converters) using BD-pTau217 (orange), eMTBR-tau (blue) and their combination (purple). Biomarkers were measured at the participant’s preceding assessment. All models were adjusted for age, sex, eGFR, BMI and assay plate. Curves were generated from held-out predictions using stratified threefold cross-validation of ridge logistic regression models. Colour-matched boxes report AUCs and 95% CIs obtained from 1,000 stratified bootstrap resamples, conditional on the held-out predictions. The grey diagonal indicates chance discrimination. Abbreviations: AD, Alzheimer’s disease; AUC, area under the curve; BMI, body mass index; CI, confidence interval; eGFR, estimated glomerular filtration rate; eMTBR-tau, endogenously cleaved microtubule-binding region tau; MCI, mild cognitive impairment; SCD, subjective cognitive decline.

### Participant characteristics and exploratory ROC analyses of conversion from MCI to AD

A total of 40 participants with MCI at their preceding assessment timepoint were assessed for progression from MCI to AD, of whom six (15.0%) converted to AD over an average follow-up of 2.0 years (**Supplementary Table S2**). At the preceding assessment, participants were on average 72.9 years old, 65.0% were female, and 35.0% were *APOE* ε4 carriers. Compared with those who remained MCI, participants who converted to AD had lower mean MMSE scores (26.7 *vs.* 28.5) and higher mean plasma pTau217 concentrations (1.13 *vs.* 0.59 pg/mL) at the preceding assessment. Importantly, all six participants that converted to AD were classified as md-aMCI at their preceding assessment. Positive plasma pTau217 status was observed in five of six converters (83.3%), compared with 11 of 34 participants who remained MCI (32.4%). *APOE* ε4 carrier status was also more frequent among converters (66.7% *vs.* 29.4%).

Among 40 participants with MCI, six converted to AD over a mean follow-up of 2.0 years (**Figure 4B**). Following adjustment for age, sex, eGFR, BMI and assay plate, cross-validated AUCs were 0.882 (95% CI, 0.716 to 1.000) for BD-pTau217, 0.936 (95% CI, 0.794 to 1.000) for eMTBR-tau and 0.922 (95% CI, 0.765 to 1.000) for the model combining both BD-pTau217 and eMTBR-tau. At a fixed predicted-probability threshold of 0.50, all three models correctly classified all 34 non-converters (specificity, 100%). However, the BD-pTau217 model identified none of the six converters (sensitivity, 0%; F1, 0), while the eMTBR-tau model identified two (sensitivity, 33.3%; F1, 0.50) and the combined model identified three (sensitivity, 50.0%; F1, 0.67).

## DISCUSSION

Using the NULISAseq™ Neuro 220 panel, we characterised plasma eMTBR-tau alongside phosphorylated tau species and other biomarkers associated with neuropathology, neurodegeneration and neuroinflammation in a brain health and memory clinic cohort. The study’s contribution lies in its detailed comparisons across clinical diagnostic groups and MCI subtypes, together with exploratory analyses of subsequent clinical progression and conversion. Here, we show that plasma biomarker differences between SCD, MCI and AD were dominated by tau biomarkers, with eMTBR-tau showing the largest fold increases in MCI compared with SCD and in AD compared with both SCD and MCI. In exploratory analyses, plasma eMTBR-tau levels at the preceding assessment showed numerically higher cross-validated discrimination of MCI-to-AD conversion over approximately two years than BD-pTau217 (AUC, 0.936 *vs*. 0.882).

Our findings extend the recent seminal work of Horie, Salvadó and colleagues^3^, who demonstrated that plasma eMTBR-tau243 measured using mass spectrometry was elevated in MCI and further elevated in AD, with strong associations with tau-PET burden and cognitive impairment^3^. Here, we confirm that eMTBR-tau is significantly elevated in MCI and AD in comparison to SCD in our cohort, characterised through comprehensive specialist and neuropsychological assessment within a multidisciplinary brain health and memory clinic that is focused on earlier stage assessment and risk reduction. Notably, we observed a concurrent significant increase of multiple phosphorylated tau species in MCI and AD, particularly BD-pTauT212, BD-pTau217 and pTau212 in MCI compared with SCD, and BD-pTauT212, BD-pTau217 and BD-pTau181 in AD compared with SCD. Moreover, complementing these clinical comparisons, eMTBR-tau abundance was 4.42-fold higher in pTau217-positive than pTau217-negative participants, further supporting its association with AD-related pathology. Indeed, *APOE* ε4 carriers showed higher abundance of several phosphorylated tau species in both SCD and MCI, and lower Aβ42 abundance in MCI, further linking genetic susceptibility to AD. Taken together, our findings reinforce a tau-dominant plasma biomarker pattern across clinical and biomarker-defined groups and highlight plasma eMTBR-tau as a promising biomarker for distinguishing clinical stages of the AD continuum.

A distinctive aspect of our study, enabled by comprehensive neuropsychological assessment, was the comparison of amnestic and non-amnestic MCI subtypes. Notably, plasma eMTBR-tau abundance was approximately 60% lower in sd-naMCI than in md-aMCI, alongside lower abundance of 12 other plasma biomarkers known to be associated with AD, predominantly phosphorylated tau species. These contrasts highlight two important points. First, MCI is heterogeneous, and combining MCI subtypes may obscure AD-associated biomarker signals that are more prominent in md-aMCI and potentially more relevant to subsequent AD progression. Second, the less pronounced AD-associated plasma tau profile in sd-naMCI supports longitudinal clinical evidence suggesting that non-amnestic MCI may progress to non-AD dementia, including dementia with Lewy bodies^20^ or frontotemporal dementia^21^. Nevertheless, it is acknowledged that future comparisons with age-matched cognitively unimpaired controls, alongside independent assessment of AD pathology, are warranted to clarify the biological significance of these subtype differences.

Another unique aspect of our study was the exploratory assessment of whether plasma BD-pTau217 and eMTBR-tau levels at the preceding assessment discriminated subsequent progression from SCD-to-MCI and conversion from MCI-to-AD over approximately two years of follow-up. Among 40 participants with MCI at their preceding assessment timepoint, six converted to AD. In this subsample, the covariate-adjusted model containing eMTBR-tau alone yielded the highest individual AUC (0.936) compared with 0.882 for BD-pTau217. Combining both BD-pTau217 and eMTBR-tau did not improve the AUC point estimate over eMTBR-tau alone, although the combined adjusted model yielded an AUC of 0.922. Notably, all six converters had md-aMCI at their preceding assessment, reinforcing this subtype as a clinically important risk phenotype, consistent with evidence that multidomain amnestic impairment carries particularly high dementia risk^22^. These exploratory findings suggest that eMTBR-tau may be a stronger predictor of MCI-to-AD progression than BD-pTau217, warranting further validation in larger independent cohorts to establish its comparative performance and potential value for risk stratification, clinical monitoring or trial enrichment^23^. Conversely, the adjusted BD-pTau217 and eMTBR-tau models showed limited discrimination of SCD-to-MCI progression (AUCs, 0.48 and 0.58, respectively), with their combination performing close to chance (AUC, 0.49). Although both exploratory ROC analyses require cautious interpretation given the small sample sizes, these findings suggest that certain markers may be more informative for discriminating progression from MCI-to-AD than earliest clinical transitions from SCD-to-MCI. It is also acknowledged, as commented on previously, the weaker discrimination of SCD-to-MCI progression may partly reflect the inclusion of naMCI subtypes, potentially diluting AD-associated biomarker signals from participants most likely to progress to AD. Consistent with this interpretation, none of the participants with SCD at baseline met criteria for md-aMCI at their two-year follow-up assessment.

### Limitations

Several limitations warrant discussion. First, while exploratory, only six participants converted from MCI-to-AD over approximately two years follow-up, limiting precision and increasing the risk of model overfitting^24^. Our exploratory progression findings should therefore be interpreted as hypothesis-generating, and be validated in independent cohorts. Second, in the absence of tau-PET data, we were unable to establish whether NULISA eMTBR-tau directly reflected cerebral tau burden or biological Braak stage. Third, while Alamar Biosciences have developed the world’s first multiplexed immunoassay to measure eMTBR-tau, analytical equivalence with the eMTBR-tau243 isoform measured using mass-spectrometry in the study by Horie, Salvadó and colleagues^3^ has not yet been established. Last, while inclusion of routine participants attending the HBA clinic is a strength, the clinic operates within a hybrid university/research setting, and as a result predominantly receives GP referrals from help-seeking participants with recent cognitive and/or mood symptoms^25^. This referral profile may limit generalisability to other memory clinics receiving participants through primary or tertiary care, where age is typically greater and where clinical severity and underlying AD prevalence may differ.

### Conclusions

Our findings reinforce a tau-dominant plasma biomarker pattern across clinical and biomarker-defined groups along the AD continuum, with eMTBR-tau showing prominent group differences. Higher plasma eMTBR-tau levels at the preceding assessment best discriminated MCI-to-AD conversion. Independent validation is now needed to determine whether NULISA-measured eMTBR-tau can inform risk stratification, guide clinical monitoring and support trial enrichment.

## Data sharing statement

The de-identified data that support the findings of this study are available from the corresponding author upon reasonable request, subject to appropriate data-sharing agreements and compliance with the conditions of the study’s ethical approvals.

## Acknowledgements

The authors would like to thank all participants and clinical staff of the Healthy Brain Ageing Program.

---

**Supplementary Figure S1.**
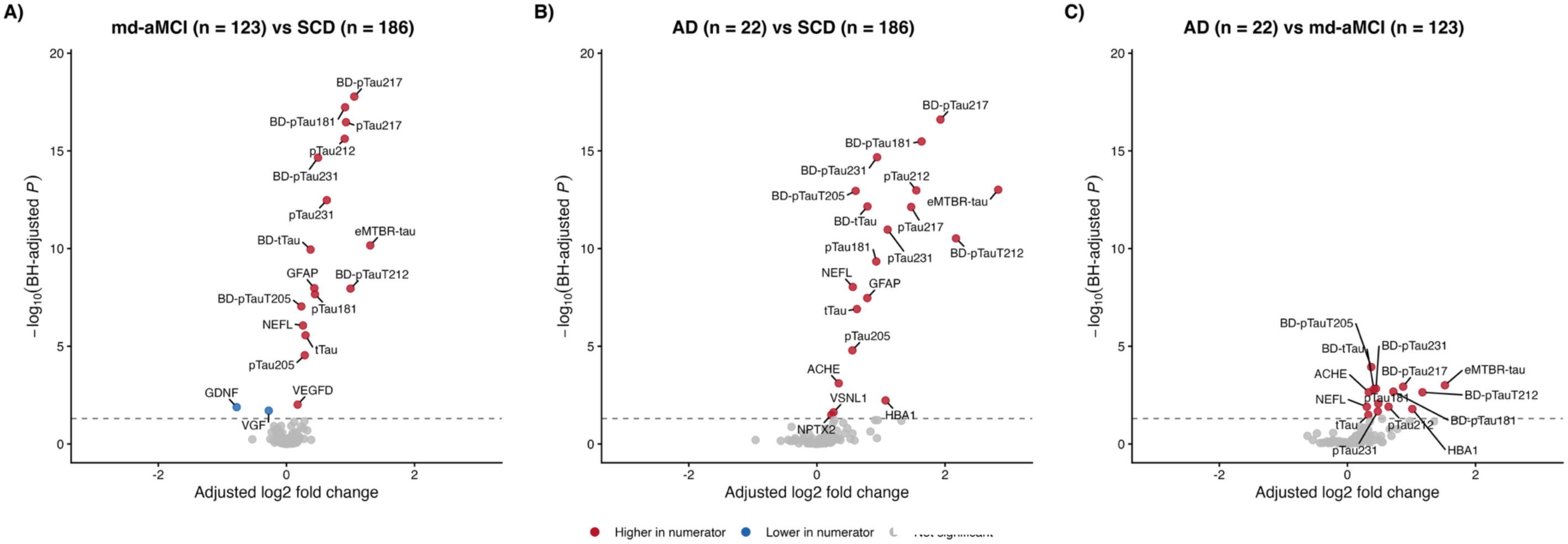
NULISAseq™ Neuro 220 panel biomarker differences across SCD, multidomain amnestic MCI and AD Volcano plots compare **A)** multidomain amnestic MCI (md-aMCI; n=123) with SCD (n=186**), B)** AD (n=22) with SCD, and **C)** AD with md-aMCI. Models were fitted to this restricted three-group sample, excluding other MCI subtypes. Analyses used one sample per participant from the latest eligible assessment and empirical Bayes moderated linear models adjusted for age, sex, *APOE* ε4 carrier status, eGFR, BMI and assay plate. P values were adjusted across tested biomarkers separately within each comparison. Each point represents a biomarker. The x-axis shows the adjusted log₂ fold change and the y-axis shows −log₁₀ of the Benjamini-Hochberg-adjusted P value. Positive values indicate higher abundance in the first named group. Red and blue points indicate significantly higher and lower abundance, respectively; grey points indicate no significant difference. The dashed horizontal line marks FDR=0.05, with significance defined as FDR < 0.05. Significantly different biomarkers are labelled. Abbreviations: AD, Alzheimer’s disease; *APOE*, apolipoprotein E; BMI, body mass index; eGFR, estimated glomerular filtration rate; FDR, false discovery rate; MCI, mild cognitive impairment; SCD, subjective cognitive decline.

**Supplementary Figure S2.**
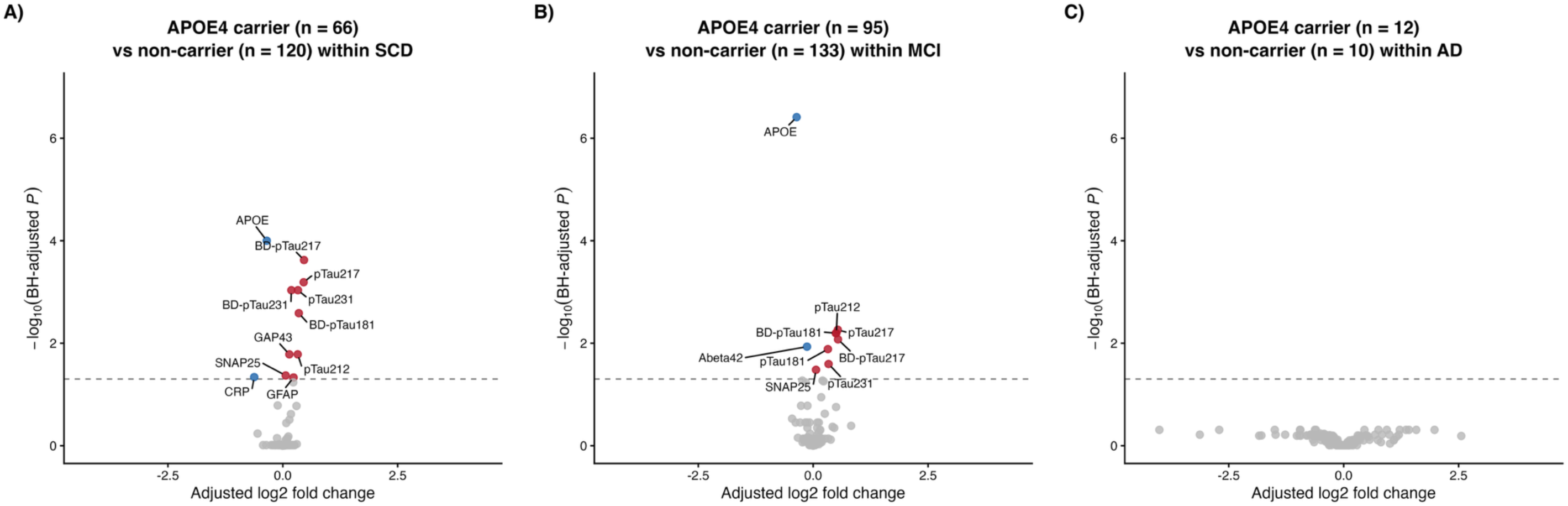
NULISAseq^TM^ Neuro 220 panel biomarker differences between *APOE* ε4 carriers and non-carriers across the AD continuum Volcano plots compare **A)** *APOE* ε4 carriers with non-carriers within SCD (66 *vs.* 120 participants), **B)** MCI (95 *vs.* 133 participants), and **C)** AD (12 *vs.* 10 participants). Analyses used one sample per participant from the latest eligible assessment and empirical Bayes moderated linear models adjusted for age, sex, eGFR, BMI and assay plate. *APOE* ε4 carrier status was the exposure and was not included as an additional covariate. P values were adjusted across tested biomarkers separately within each comparison. Each point represents a biomarker. The x-axis shows the adjusted log₂ fold change and the y-axis shows −log₁₀ of the Benjamini-Hochberg-adjusted P value. Positive values indicate higher abundance in *APOE* ε4 carriers. Red and blue points indicate significantly higher and lower abundance, respectively; grey points indicate no significant difference. The dashed horizontal line marks FDR=0.05, with significance defined as FDR < 0.05. Significantly different biomarkers are labelled. Abbreviations: AD, Alzheimer’s disease; *APOE*, apolipoprotein E; BMI, body mass index; eGFR, estimated glomerular filtration rate; FDR, false discovery rate; MCI, mild cognitive impairment; SCD, subjective cognitive decline.

**Supplementary Table S1.** Characteristics of participants assessed for progression from SCD to MCI.

| Characteristic | All ROC participants | Remained SCD at last visit | Converted to MCI by last visit |
| --- | --- | --- | --- |
| Participants, n | 106 | 87 | 19 |
| Follow-up time (years) | 2.0 (0.4) | 2.0 (0.5) | 2.0 (0.4) |
| Age (years) | 67.1 (7.6) | 67.0 (7.5) | 67.6 (8.1) |
| Sex, female | 75 (70.8%) | 58 (66.7%) | 17 (89.5%) |
| Education (years) | 14.8 (2.8) | 14.9 (2.9) | 14.5 (2.1) |
| MMSE at preceding assessment | 29.4 (0.8) | 29.3 (0.9) | 29.7 (0.5) |
| BMI (kg/m<sup>2</sup>) | 26.6 (4.5) | 26.6 (4.6) | 26.5 (4.2) |
| *APOE* ε4 carriers | 37 (34.9%) | 31 (35.6%) | 6 (31.6%) |
| Plasma pTau217 (pg/mL) | 0.27 (0.191) | 0.27 (0.194) | 0.26 (0.182) |
| Plasma pTau217 status<sup>1</sup> |  |  |  |
| Negative | 88 (83.0%) | 73 (83.9%) | 15 (78.9%) |
| Indeterminate | 12 (11.3%) | 10 (11.5%) | 2 (10.5%) |
| Positive | 6 (5.7%) | 4 (4.6%) | 2 (10.5%) |

Continuous values are mean (SD); categorical values are count (%) among observed values. Missing data are summarised in Supplementary Table S3
Abbreviations: MMSE, Mini-Mental State Examination; eGFR, estimated glomerular filtration rate; *APOE*, apolipoprotein E; MCI, mild cognitive impairment; SCD, subjective cognitive decline; AD, Alzheimer's disease
<sup>1</sup>Study pTau217 thresholds: negative <0.4; indeterminate 0.40 to <0.64; positive ≥0.64 pg/mL, using the combined measured/converted concentration. Thresholds are based on the SIMOA ALZpath references reported by Ashton et al (2024).

**Supplementary Table S2.**
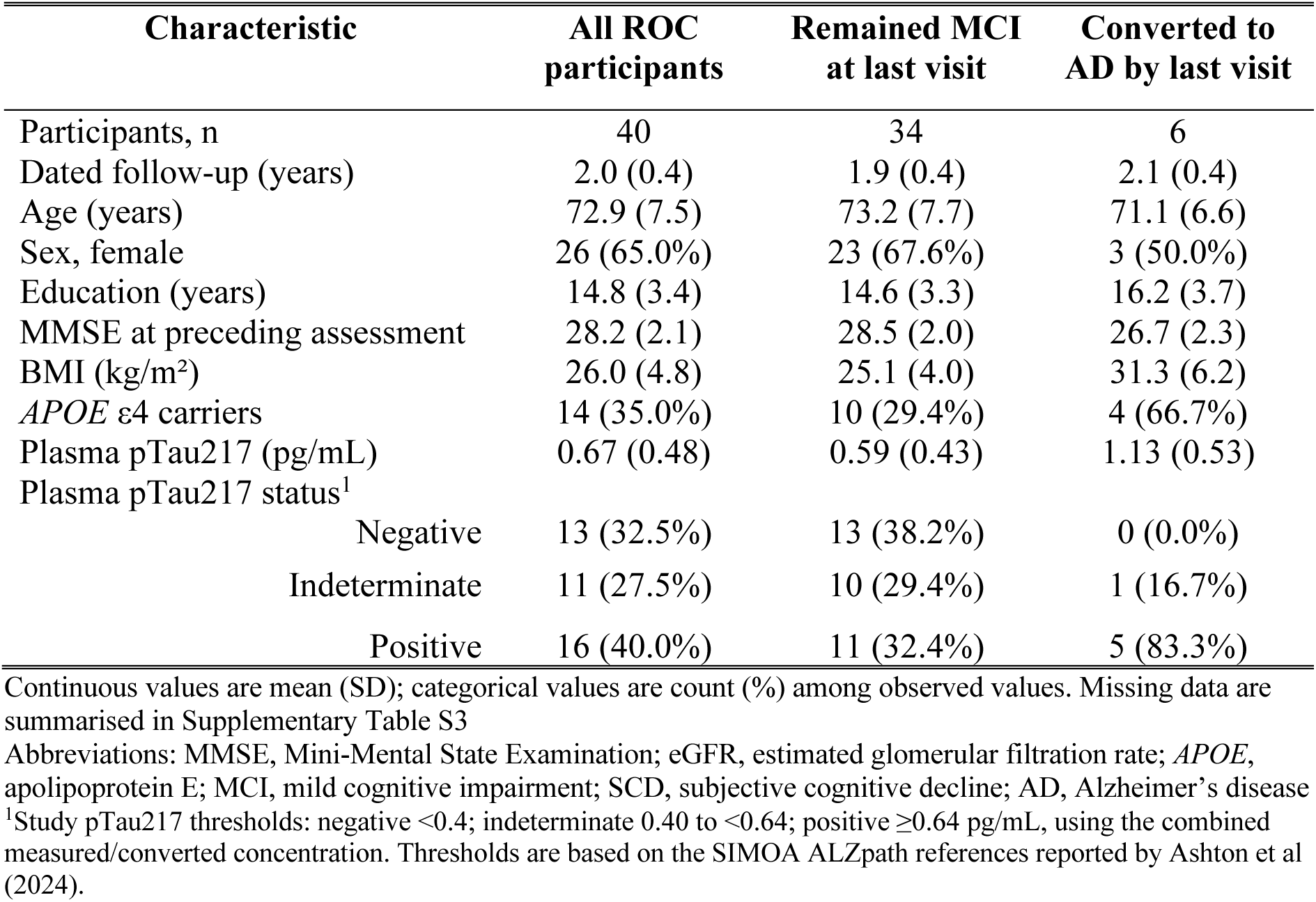
Characteristics of participants assessed for conversion from MCI to AD.

| Characteristic | All ROC participants | Remained MCI at last visit | Converted to AD by last visit |
| --- | --- | --- | --- |
| Participants, n | 40 | 34 | 6 |
| Dated follow-up (years) | 2.0 (0.4) | 1.9 (0.4) | 2.1 (0.4) |
| Age (years) | 72.9 (7.5) | 73.2 (7.7) | 71.1 (6.6) |
| Sex, female | 26 (65.0%) | 23 (67.6%) | 3 (50.0%) |
| Education (years) | 14.8 (3.4) | 14.6 (3.3) | 16.2 (3.7) |
| MMSE at preceding assessment | 28.2 (2.1) | 28.5 (2.0) | 26.7 (2.3) |
| BMI (kg/m<sup>2</sup>) | 26.0 (4.8) | 25.1 (4.0) | 31.3 (6.2) |
| *APOE* ε4 carriers | 14 (35.0%) | 10 (29.4%) | 4 (66.7%) |
| Plasma pTau217 (pg/mL) | 0.67 (0.48) | 0.59 (0.43) | 1.13 (0.53) |
| Plasma pTau217 status<sup>1</sup> |  |  |  |
| Negative | 13 (32.5%) | 13 (38.2%) | 0 (0.0%) |
| Indeterminate | 11 (27.5%) | 10 (29.4%) | 1 (16.7%) |
| Positive | 16 (40.0%) | 11 (32.4%) | 5 (83.3%) |

Continuous values are mean (SD); categorical values are count (%) among observed values. Missing data are summarised in Supplementary Table S3
Abbreviations: MMSE, Mini-Mental State Examination; eGFR, estimated glomerular filtration rate; *APOE*, apolipoprotein E; MCI, mild cognitive impairment; SCD, subjective cognitive decline; AD, Alzheimer's disease
<sup>1</sup>Study pTau217 thresholds: negative <0.4; indeterminate 0.40 to <0.64; positive ≥0.64 pg/mL, using the combined measured/converted concentration. Thresholds are based on the SIMOA ALZpath references reported by Ashton et al (2024).

**Supplementary Table S3.** Missing data across tables 1, S1 and S2.

| Table | Characteristic | Group | Participants | Observed | Missing, n (%) |
| --- | --- | --- | --- | --- | --- |
| Table 1 | Education (years) | Overall | 436 | 428 | 8 (1.8%) |
| Table 1 | Education (years) | SCD | 186 | 182 | 4 (2.2%) |
| Table 1 | Education (years) | MCI | 228 | 224 | 4 (1.8%) |
| Table 1 | Education (years) | AD | 22 | 22 | 0 (0.0%) |
| Table 1 | MMSE | Overall | 436 | 424 | 12 (2.8%) |
| Table 1 | MMSE | SCD | 186 | 186 | 0 (0.0%) |
| Table 1 | MMSE | MCI | 228 | 219 | 9 (3.9%) |
| Table 1 | MMSE | AD | 22 | 19 | 3 (13.6%) |
| Table 1 | Plasma pTau217 (pg/mL) | Overall | 436 | 434 | 2 (0.5%) |
| Table 1 | Plasma pTau217 (pg/mL) | SCD | 186 | 186 | 0 (0.0%) |
| Table 1 | Plasma pTau217 (pg/mL) | MCI | 228 | 228 | 0 (0.0%) |
| Table 1 | Plasma pTau217 (pg/mL) | AD | 22 | 20 | 2 (9.1%) |
| Table 1 | Plasma pTau217 status | Overall | 436 | 434 | 2 (0.5%) |
| Table 1 | Plasma pTau217 status | SCD | 186 | 186 | 0 (0.0%) |
| Table 1 | Plasma pTau217 status | MCI | 228 | 228 | 0 (0.0%) |
| Table 1 | Plasma pTau217 status | AD | 22 | 20 | 2 (9.1%) |
| Supplementary Table S1 | MMSE at preceding assessment | All SCD-to-MCI ROC participants | 106 | 67 | 39 (36.8%) |
| Supplementary Table S1 | MMSE at preceding assessment | Remained SCD | 87 | 52 | 35 (40.2%) |
| Supplementary Table S1 | MMSE at preceding assessment | Progressed to MCI | 19 | 15 | 4 (21.1%) |
| Supplementary Table S2 | Education (years) | All MCI-to-AD ROC participants | 40 | 39 | 1 (2.5%) |
| Supplementary Table S2 | Education (years) | Remained MCI | 34 | 33 | 1 (2.9%) |
| Supplementary Table S2 | Education (years) | Converted to AD | 6 | 6 | 0 (0.0%) |
| Supplementary Table S2 | MMSE at preceding assessment | All MCI-to-AD ROC participants | 40 | 21 | 19 (47.5%) |
| Supplementary Table S2 | MMSE at preceding assessment | Remained MCI | 34 | 18 | 16 (47.1%) |
| Supplementary Table S2 | MMSE at preceding assessment | Converted to AD | 6 | 3 | 3 (50.0%) |

Only characteristics with missing values are shown. Missing percentages use the total number of participants in each table column; Abbreviations: MMSE, Mini-Mental State Examination

